# Joint modelling of extremely longitudinal measurements and competing survival outcomes in HIV-infected patients

**DOI:** 10.1101/2025.03.12.25323864

**Authors:** Yifan Tang, Hongfei Zhu, Kai Wang, Yifan Chen, Ruwanthi Kolamunnage-Dona, Wei Cheng, Ying Wang, Chengxiu Ling, Chengliang Chai, Na He

## Abstract

**Background:** The CD4 T-lymphocyte (CD4) count is a critical biomarker for HIV disease progression and immune health, that is essential for guiding treatment decisions. However, quantitative assessment of the impact of rapid CD4 decline on AIDS progression and pre-AIDS death remains unclear.

**Methods:** This study includes 11,647 HIV-positive patients from the Zhejiang Provincial Center for Disease Control and Prevention (CDC) from 2005 to 2017. The longitudinal trajectory of extremely low CD4 counts is captured by a generalized Pareto (GP) model, jointly analyzed with two competing events (AIDS progression and pre-AIDS death) via Weibull spatial survival models. The survival submodels are linked through a shared linear predictor in the GP submodel, providing insights into the effect of extremely low CD4 counts on competing outcomes. The model is implemented using the efficient R-INLA (integrated nested Laplace approximations) approach, with predictive performance assessed using survival Receiver Operating Characteristic (ROC) curves and integrated Area Under the Curve (iAUC) values.

**Results:** Male individuals, those with homosexual transmission, older age at diagnosis, and extended delays in starting initial antiviral therapy (ART) are more likely to experience sharper reduction in CD4 counts. This decline, along with female gender and late ART initiation, increases the risk of both AIDS progression and pre-AIDS death, with a more pronounced effect on the former. The considerate spatial survival frailty structure with the shared GP longitudinal model for the sharp decline of CD4 counts enhances the predictive accuracy for both outcomes in comparison with the Gaussian longitudinal submodel for CD4 counts (iAUC: 0.9183 vs 0.7688 (AIDS progression) and 0.8901 vs 0.6634 (pre-AIDS death)).

**Conclusion:** Our study confirms that demographic factors, route of infection, diagnosis related factors, and extremely low CD4 counts have a significant impact on AIDS progression and pre-AIDS death. These findings underscore the importance of developing effective, evidence-based strategies to mitigate the risk of HIV/AIDS.

## 1 Introduction

Acquired Immune Deficiency Syndrome (AIDS) is an incurable disease caused by the Human Immunodeficiency Virus (HIV) (Farhadian et al., 2021). By the end of 2022, approximately 39 million people were living with HIV worldwide, with 1.3 million new cases and 630,000 deaths from HIV-related causes in 2022 (WHO, 2023), while China reported 1.223 million cases of HIV/AIDS, with 107,000 new cases (China CDC, 2023; He, 2021). The increasing AIDS risks or death among HIV-infected individuals are severe in regions with limited economic development and high population mobility, such as sub-Saharan Africa, Latin America (Coelho et al., 2021), South-East Asia, Western Pacific regions (Gilmour et al., 2023), and Korea (Park et al., 2022).

Most biomarkers for the progression of AIDS are related to the CD4 T-lymphocyte (CD4) cell count (Lembas et al., 2022; Lin et al., 2019). It is primarily targeted by HIV, leading to its massive destruction and depletion, which progressively impairs the body’s immune function. The amount of CD4 cells is considered a critical biological marker for understanding the disease progression and guiding the initiation of antiretroviral therapy (ART), among other key clinical management decisions (Karim and Karim, 2010). For instance, according to the World Health Organization (WHO), one of the signs of AIDS progression and a deadly AIDS status is a CD4 count falling below 200 and 150 cells*/*mm^3^ (Morpeth et al., 2007).

Numerous modelling approaches have been established to quantify the risk progression of HIV-infected individuals and its critical indicators. AIDS progression and pre-AIDS death (primarily due to suicide), as two major outcomes among HIV-infected individuals (Park et al., 2022), have been jointly analyzed with the longitudinal CD4 profiles in the Bayesian framework (Anglaret et al., 2012; Momenyan, 2021), accounting for the heterogeneous (patient-specific) CD4 variability (Andrinopoulou et al., 2017; Mchunu et al., 2022) and spatial survival frailty effects (Martins et al., 2016; Momenyan, 2021). The joint modeling method offers advantages like bias reduction and improvement estimation efficiency, by incorporating the dependence between longitudinal and survival data (Dessiso and Goshu, 2017; Martins et al., 2016) through the use of a shared parametric model (Mchunu et al., 2022). This approach avoids the spurious results often produced in traditional Cox/Weibull hazard models with time-varying longitudinal profiles, as its association with informative drop-out is neglected, as seen in studies of cancer and AIDS (Campbell et al., 2019; Henderson et al., 2000; Rizopoulos, 2012). For broader applications of joint models in longitudinal and survival data, we refer to relevant work on diabetes progression and colorectal cancer mortality (Dennis et al., 2018; Król et al., 2016).

Note that most joint models aforementioned rely on mean-based Gaussian variability for longitudinal CD4 counts data (Hickey et al., 2018; Thomadakis et al., 2024), which are exposed to substantial measurement errors, resulting in its skewness and heavy tail even after standardization (Zhang and Huang, 2021). Consequently, some alternative approaches for such longitudinal biomarkers were proposed with evidenced good performance, including robust quantile regression (Zhang and Huang, 2021) for viral load amount in HIV-positive patients, and multivariate skew *t* distributions for chronic kidney disease (Ferede et al., 2022). This study aims to establish a comparable Bayesian joint model for the extreme features of longitudinal CD4 profiles and two competing survival outcomes (AIDS progression and pre-AIDS death), incorporating both subjects’ characteristics (age, education level, marital status, contact pattern) (Farhadian et al., 2021) and spatial frailty effects.

Although the extreme values of longitudinal biomarkers (CD4 counts, viral loads) are clearly relevant for studying the progression of HIV risks, no research comprehensively analyzes longitudinal extreme CD4 trajectories either separately or jointly with infection progression. This study aims to bridge this gap by jointly analyzing extreme reductions in CD4 counts and competing survival outcomes. First, we employed the generalized Pareto (GP) model for the downward excess of longitudinal CD4 counts, following the extreme statistical modeling methodology proposed recently for longitudinal data (Spearing et al., 2023), see e.g., Cai et al. (2023); Pan et al. (2024) for the applications of GP models studying air pollution and infectious diseases. Subsequently, the linear predictor of the GP model, consisting of fixed effects associated with demographic characteristics and within-subject temporal effects, will be shared to the Weibull competing hazards models for infection progression. The model is implemented using the efficient R-INLA approach (Niekerk et al., 2021; Rustand et al., 2024) with model performance evaluated using the Deviance Information Criterion (DIC) (Wang et al., 2023a). Finally, we demonstrate the predictive performance of our competing survival models, with the use of survival Receiver Operating Characteristics (ROC) curves and integrated Area Under the Curve (iAUC) values (Heagerty and Zheng, 2005; Saha-Chaudhuri and Heagerty, 2013; Uno et al., 2007), in comparison with those with shared Gaussian longitudinal submodel (Momenyan, 2021).

The remainder of this paper is organized as follows. In Section 2, we present our HIV infection data and covariates. In Section 3, we establish the joint Bayesian models for sharp reduction of longitudinal CD4 profiles and two competing outcomes (AIDS progression and pre-AIDS death). Section 4 is devoted to model results and evaluations. We give an extensive discussion and conclusion in Sections 5 and 6, respectively.

## 2 Data

### 2.1 CD4 counts and time to events

We collected data from the HIV/AIDS case reporting information system and the national HIV treatment subdatabase of Zhejiang Province on 21,711 HIV-positive patients from 89 districts and counties in Zhejiang Province from January 2005 to December 2017. The final analysis sample size was 11,647. Exclusion criteria included: 6,494 individuals with pre-existing AIDS based on the diagnostic criteria from the Chinese Guidelines for Diagnosis and Treatment of AIDS (diagnosed with HIV infection and CD4 counts < 200 cells*/*mm^3^) (Li et al., 2024), 2,982 individuals with fewer than three recorded CD4 counts, 27 individuals with a marital status of “other”, and 561 individuals with routes of infection classified as “homosexual/heterosexual transmission”.

In our analysis of longitudinal trajectory, we primarily focus on CD4 counts measured over time since the HIV diagnosis. A total of 102,842 measurements are recorded from 11,647 individuals, with an average of 8.83 measurements per patient, and each patient is monitored between 3 and 38 times during the follow-up period. The longitudinal trajectories of CD4 measurements among 50 randomly selected individuals are shown in Figure 1(a), with the raw counts exhibiting a large standard deviation of 187.52 and a skewness of 1.12, indicating substantial variation in baseline levels. Log-transformed CD4 counts were used for subsequent analysis.

**Figure 1:**
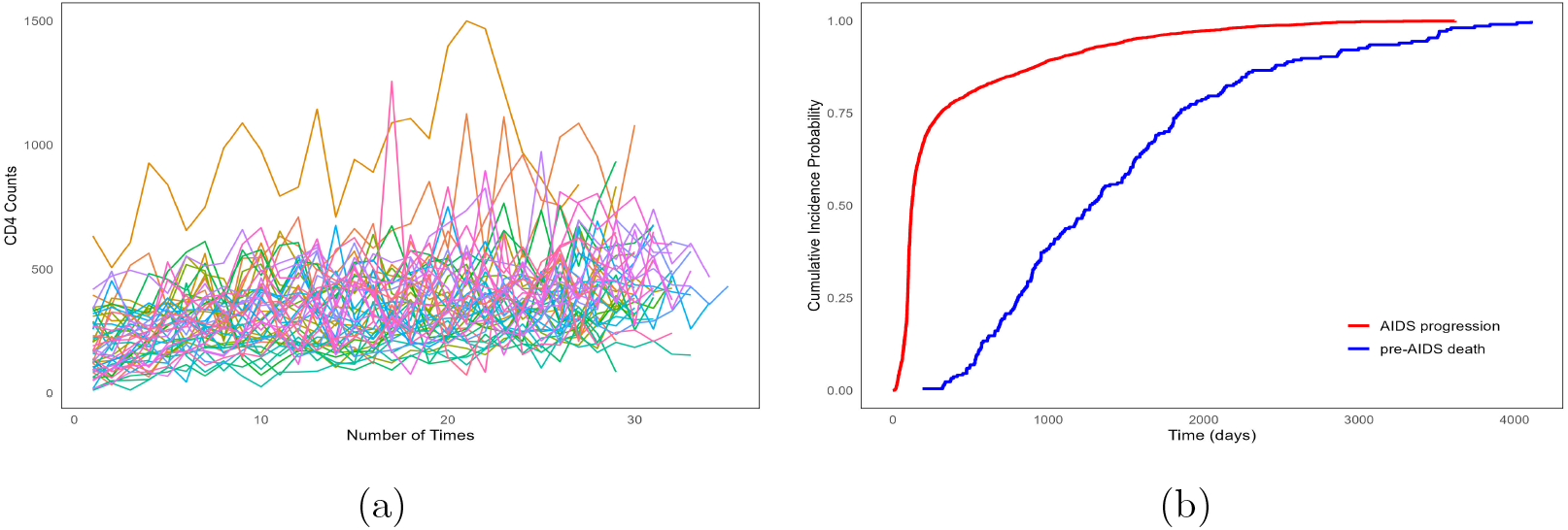
Longitudinal trajectories of CD4 measurements (a) and the cumulative incidence functions for AIDS progression and pre-AIDS death (b).

As for the health outcomes, we examined the risk of AIDS and pre-AIDS death among HIV-infected individuals. Specifically, AIDS progression was defined as the event in which an individual’s CD4 count dropped below 200 cells*/*mm^3^ during follow-up measurements, marking the onset of AIDS. Pre-AIDS death, on the other hand, referred to individuals who died before reaching a CD4 count below 200 cells*/*mm^3^ without progressing to AIDS. Of the 11,647 individuals, 966 (8.29%) progressed to AIDS, 89 (0.76%) died before reaching AIDS, and 10,593 (90.95%) were censored (i.e., lost to follow-up or do not experience either outcome by December 30, 2017). Furthermore, we see a consistently higher risk of AIDS progression throughout the follow-up period in Figure 1(b).

### 2.2 Explanatory variables

The explanatory variables for this study are listed in Table 1, including (1) gender (male, female); (2) marital status (single, married); (3) route of infection (homosexual, heterosexual); (4) age at HIV-infected diagnosis in years, (5) the treatment delay, measured as the number of days from the HIV-positive diagnosis to the initiation of ART. Among the 11,647, the majority are male (9,616, 82.56%) and single (6,749, 57.95%), the median age at diagnosis is 33 years, and the average treatment delay is 281 days. Table 1 reveals that gender, marital status, age at diagnosis, and treatment delay are significantly associated with AIDS progression or pre-AIDS death (*p <* 0.05), while no significant difference is observed for the route of infection.

**Table 1:** The descriptive analysis of characteristics of HIV-infected patients.

| Explanatory Variables | Frequency (%) | Log-rank | *p*-value<sup>b</sup> |
| --- | --- | --- | --- |
| **Gender** |  |  |  |
| Male | 9616 (82.56) | 5.249 | 0.022 |
| Female | 2031 (17.44) |  |  |
| **Marital status** |  |  |  |
| Single<sup>a</sup> | 6749 (57.95) | 4.659 | 0.031 |
| Married | 4898 (42.05) |  |  |
| **Route of infection** |  |  |  |
| Homosexual transmission | 5895 (50.61) | 0.438 | 0.508 |
| Heterosexual transmission | 5752 (49.39) |  |  |
| **Age at diagnosis (year)** |  |  |  |
| 20– | 430 (3.68) | 6845 | <0.01 |
| 21 to 30 | 4690 (40.17) |  |  |
| 31 to 40 | 2789 (23.89) |  |  |
| 41 to 50 | 1878 (16.09) |  |  |
| 51+ | 1860 (15.93) |  |  |
| **Treatment delay (day)** |  |  |  |
| 20– | 3371 (28.94) | 1003 | <0.01 |
| 21 to 40 | 2180 (18.72) |  |  |
| 41 to 80 | 1373 (11.79) |  |  |
| 81 to 160 | 941 (8.08) |  |  |
| 161 to 320 | 994 (8.53) |  |  |
| 321+ | 2788 (23.94) |  |  |

a. The single status includes unmarried, divorced, and widowed.
b. The disparities in time to AIDS progression or pre-AIDS death among various groups are assessed using the log-rank test, with statistical significance determined at a 5% level in two-tailed tests.

This study is conducted in accordance with the ethical principles of the Declaration of Helsinki. It is approved by the Institutional Review Board of the School of Public Health, Fudan University (#2014-03-0497).

## 3 Joint model formulation

The joint model consists of two submodels: a GP submodel for longitudinal trajectories of extremely low CD4 counts and a competing risks Weibull survival submodel for the two competing events (AIDS progression and pre-AIDS death). These submodels are connected by sharing the linear predictor from the longitudinal submodel with the survival submodel, enabling the longitudinal assessment of the effect of extremely low CD4 counts on both competing outcomes (Niekerk et al., 2021). The censoring mechanism in the survival process is presumed to be non-informative conditionally on the longitudinal CD4 profiles.

### 3.1 Longitudinal submodel with peaks-over-threshold approach

Let *y*(*t*) be the logarithmic transformation of the CD4 counts measured at day *t* since the HIV-infected, ranging in [1, 5118]. For the extremely low CD4 counts, we consider the downward excess modelled by GP distribution (Cai et al., 2023; Pan et al., 2024):

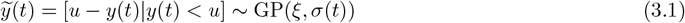

for a small threshold *u* indicating extremely low CD4 counts. Here *ξ* is the shape parameter, which is held as constant, while the scale parameter *σ*(*t*) can be reparameterized in terms of 100*α*% quantile (*μ*(*t*)) via *μ*(*t*) = *σ*(*t*) *·* ((1 *− α*)^*−ξ*^ *−* 1)*/ξ* (Opitz et al., 2018). A detailed explanation of this method is provided in Appendix A.1, and it has also been applied to model the downward excess of birth rates in the U.S. (Wang et al., 2023b) and elite swimming (Spearing et al., 2023).

In this study, we propose a linear mixed model for the 60% quantile of the downward excess CD4 specified in Eq.(3.1) with log-link function. Specifically,

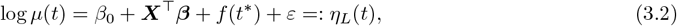

where *β*_0_ is the intercept, ***β*** is the fixed effect associated with baseline predictors ***X*** introduced in Section 2.2 consisting of gender, marital status, route of infection, age of HIV diagnosis, and the delay of treatment since HIV-positive diagnosis. In addition, the Gaussian measurement error *ε* plays the role of individualized random intercept. The temporal random effect *f* (*t*^*\**^), *t*^*\**^ = 1, 2, …, 50 is a second-order random walk (RW2) model that facilitates a smoothing variation in time, with independent second-order increments (Rue and Held, 2005),

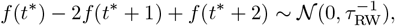

where *t*^*\**^ ranges from 1 to 50, standing for the 50 equal-spaced period containing the original time interval [1, 5118]. The length of each period is around 102 days, aligning with the time difference between the repeated measures (usually quarter-yearly) (Li et al., 2024). The 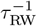 is the variance of the white noise process.

### 3.2 Survival submodels and joint model

For the survival component, let *d* = 0, 1, 2 be the event type indicator, where *d* = 0 represents the non-informative censorship, and *d* = 1, 2 denote the competing causes (event) of AIDS progression and pre-AIDS death progression, respectively. We consider Weibull survival models for the two cause-specific hazard functions, defined as 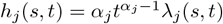, *j* = 1, 2. Each hazard function has a constant shape parameter *α*_*j*_ and a scale parameter *λ*_*j*_(*s, t*) (Niekerk et al., 2021), which varies spatially across the counties and districts (*s*) in Zhejiang province and temporally changes with survival (censoring) time *t*. This scale parameter is modelled by a log-linear component with mixed effects,

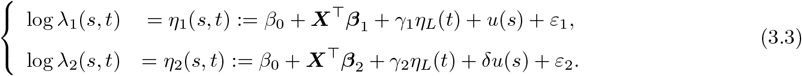

The intercepts *β*_0_ are consistent with that in the longitudinal submodel, and *ε*_1_, *ε*_2_ are the independently individualized random intercepts. The fixed effects ***β***_1_ and ***β***_2_ are associated with the same predictors ***X***, as utilized in the longitudinal submodel. To enhance model parsimony and reduce uncertainty in estimating these effects, a shared spatial random effect *u*(*s*) is employed across the survival submodels, scaled by *δ*. This random effect is constructed by the Besag-York-Mollié 2 (BYM2) model (Riebler et al., 2016), which intricately combines the structured Besag (intrinsic conditional autoregressive) random effect *u*_*\**_(*s*) with the un-structured (namely I.I.D.) random effect *v*_*\**_(*s*):

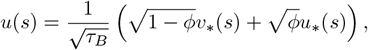

where the mixing parameter *ϕ ∈* [0, 1] serves as a spatial smoothing parameter, measuring the proportion of the marginal variance explained by the structured term *u*_*\**_(*s*).

Furthermore, to evaluate the influence of CD4 counts on the two events and estimate the parameters of both longitudinal and survival processes simultaneously, we incorporate the linear predictor *η*_*L*_(*t*) (cf. Eq.(3.2)) from the longitudinal submodel into the two survival submodels, utilizing the shared coefficients *γ*_1_ and *γ*_2_. This approach enhances model parsimony while allowing us to assess the potential similarities or differences in the effects of the longitudinal trajectory of CD4 on the two competing causes based on the signs of the sharing coefficients.

### 3.3 Priors definition

In the joint model, we set default log-gamma priors in INLA for the tail parameter *ξ* in the GP model and employ log-Gaussian priors for the shape parameter *α*_1_, *α*_2_ in the cause-specific Weibull survival models. The priors for the other parameters follow the specifications outlined by Niekerk et al. (2021). We set vague Gaussian priors for the fixed effects ***β, β***_1_, ***β***_2_ and sharing coefficients *γ*_1_, *γ*_2_, *δ*, and use log-gamma prior for the Gaussian precision *τ*_*ε*_ for *ε, ε*_1_, *ε*_2_. For the spatial and temporal model parameters, we employ the penalized complexity (PC) priors (Simpson et al., 2017) that penalize the complex models to avoid overfitness. We set Prob(*τ*_RW_ *<* 1) = 0.01 for the RW model, which means the probability of the standard deviation 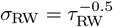 larger than 1 is very low. For the BYM2 model, we use Prob(*τ*_BYM_ *<* 1.6) = 0.01 to penalize model with large variance and Prob(*ϕ >* 0.1) = 0.80.

The sensitivity analysis utilized default priors in INLA. The results confirm the robustness of all the priors we defined, with consistent posterior estimations of fixed effects and parameters.

### 3.4 Model evaluation, diagnosis and selection

To assess the goodness-of-fit of the joint model with spatiotemporal terms, we compare it with parsimonious models excluding either spatial *u*(*s*), temporal *f* (*t*^*\**^), or both terms. Model performance was evaluated using the Deviance Information Criterion (DIC), which accounts for both the fitness based on posterior estimates and penalization of complexity, with lower values indicating better-performing models (Wang et al., 2023a).

Additionally, to evaluate the accuracy of the survival model, we conduct a log-Gaussian submodel for comparison, which captures the longitudinal trajectories of all CD4 count records without restricting the analysis to extremely low counts, while maintaining the same competing risk structure (see Appendix A.2). We employ survival ROC curves, survival AUC and iAUC values (Heagerty and Zheng, 2005; Saha-Chaudhuri and Heagerty, 2013; Uno et al., 2007) to assess the accuracy of both survival models for each outcome.

## 4 Results

### 4.1 Model results

In the GP sub-model for the downward excess of CD4 counts (on log scale), we choose the 15% quantile *u* = 5.7 (cf. Eq.(3.1)) as the threshold, following the rule of thumb (Haan and Ferreira, 2006). Correspondingly, in terms of the original CD4 counts, the threshold translates to approximately exp(5.7) *≈* 299 cells*/*mm^3^. Accordingly, there are 57.5% (6,756 out of 11,756) HIV-patients with at least one CD4 counts record less than 299 (extremely low CD4 in the context). We found that 15% (975 out of 6,756) progressed to AIDS or died before AIDS, which is significantly higher in comparison with the progression percentage 2% (80 out of 4,991) without any low CD4 counts (*p <* 0.05). Subsequently, we investigate the GP-based longitudinal downward excess of CD4 counts jointly with the two cause-specific hazards, including AIDS progression and pre-AIDS death. The influential factors associated with the progression of HIV-infected individuals and the posterior estimates of parameters involved in the models are presented in Tables 2 and 3.

**Table 2:** Summary of posterior estimates of fixed effect in the joint model fitted by competing risks (AIDS progression and pre-AIDS death) and longitudinal measurements (CD4 counts).

| **Longitudinal endpoint: downward excess of CD4 count** |  |  |  |
| --- | --- | --- | --- |
| **Predictors** | Mean | 2.5% quantile | 97.5% quantile |
| **Gender** |  |  |  |
| Female | Reference |  |  |
| Male | 0.213 | 0.170 | 0.257 |
| **Marital status** |  |  |  |
| Single | Reference |  |  |
| Married | 0.006 | −0.033 | 0.047 |
| **Route of infection** |  |  |  |
| Heterosexual transmission | Reference |  |  |
| Homosexual transmission | 0.083 | 0.041 | 0.124 |
| **Age at diagnosis** | 0.005 | 0.003 | 0.007 |
| **Treatment delay** | 0.002 | 0.001 | 0.003 |
| **Survival endpoint 1: AIDS progression** |  |  |  |
| **Predictors** | Mean | 2.5% quantile | 97.5% quantile |
| **Gender** |  |  |  |
| Female | Reference |  |  |
| Male | −1.994 | −2.362 | −1.625 |
| **Marital status** |  |  |  |
| Single | Reference |  |  |
| Married | −0.068 | −0.421 | 0.285 |
| **Route of infection** |  |  |  |
| Heterosexual transmission | Reference |  |  |
| Homosexual transmission | −0.840 | −1.204 | −0.475 |
| **Age at diagnosis** | −0.048 | −0.060 | −0.037 |
| **Treatment delay** | −0.018 | −0.021 | −0.015 |
| **Survival endpoint 2: Pre-AIDS death** |  |  |  |
| **Predictors** | Mean | 2.5% quantile | 97.5% quantile |
| **Gender** |  |  |  |
| Female | Reference |  |  |
| Male | −1.729 | −2.324 | −1.133 |
| **Marital status** |  |  |  |
| Single | Reference |  |  |
| Married | −0.433 | −1.045 | 0.179 |
| **Route of infection** |  |  |  |
| Heterosexual transmission | Reference |  |  |
| Homosexual transmission | −0.569 | −1.282 | 0.144 |
| **Age at diagnosis** | −0.003 | −0.021 | 0.013 |
| **Treatment delay** | −0.021 | −0.032 | −0.009 |

**Table 3:** Summary of the posterior estimates of the parameters.

| Parameter | Mean | 2.5% quantile | 97.5% quantile |
| --- | --- | --- | --- |
| Tail parameter ($\xi$, in $10^{-3}$) | 1.196 | 0.984 | 1.461 |
| Shape parameter of AIDS progression ($\alpha_1$) | 2.627 | 2.584 | 2.686 |
| Shape parameter of pre-AIDS death ($\alpha_2$) | 0.797 | 0.786 | 0.810 |
| BYM2 mixing parameter ($\phi$) | 0.075 | 0.057 | 0.086 |
| Shared scale of AIDS progression ($\gamma_1$) | 7.885 | 7.770 | 8.008 |
| Shared scale of pre-AIDS death ($\gamma_2$) | 2.472 | 2.299 | 2.647 |

First, we find the extreme reduction of CD4 counts are significantly associated with gender, route of infection, age at diagnosis, and treatment delay (cf. Table 2 for the longitudinal endpoint). Specifically, males are expected to experience 1.231 (exp(0.213), 95% CI: 1.170, 1.257) times decline in CD4 counts compared to females, and individuals with homosexual transmission are 1.083 (95% CI: 1.041, 1.124) times greater than those with heterosexual transmission. Additionally, a one-year increase in age at diagnosis and a ten-day increase in the interval between diagnosis and treatment are associated with a decline CD4 count by factors of 1.005 (95% CI: 1.003, 1.007) and 1.002 (95% CI: 1.001, 1.003), respectively. This dataset didn’t show any significant difference of the declining CD4 counts between single and married HIV-infected individuals.

Second, all predictors show similar associations of AIDS progression risks to the extreme reduction of CD4 counts. Namely, except for marital status, all other predictors (gender, route of infection, age at diagnosis, and treatment delay) show significant effects on the risk of AIDS progression. Specifically, the hazard ratio AIDS progression in males is expected to be 86.38% (1 *−* exp(*−*1.994), 95% CI: 0.803, 0.893) lower for females, and for individuals with homosexual transmission, it is 56.82% (95% CI: 0.378, 0.700) lower compared to those with heterosexual transmission. Additionally, each year earlier at diagnosis and every 10 days earlier initiation of ART treatment since HIV diagnosis are associated with 4.68% (95% CI: 0.036, 0.058) and 1.78% (95% CI: 0.015, 0.021) decrease in the hazard ratio for AIDS progression, respectively.

Third, less significant predictors are identified for pre-AIDS death risks in contrast with the AIDS progression. Male mortality before AIDS progression is lower than that of females, with an 82.25% (95% CI: 0.678, 0.902) hazard ratio, and every 10 days earlier initiation of ART treatment since HIV diagnosis is associated with a 2.07% (95% CI: 0.003, 0.031) decrease in the hazard ratio of pre-AIDS death. Conversely, none of marital status, route of infection, or age at diagnosis show a significant influence on pre-AIDS death.

The posterior estimates of parameters are shown in Table 3, including the tail parameter (*ξ*) in the GP distribution, the shape parameters (*α*_1_, *α*_2_) for Weibull models, the mixing parameter (*ϕ*) for BYM2 model, and the regression coefficients (*γ*_1_, *γ*_2_) of the linear predictor in the longitudinal model shared into the Weibull models. In particular, the estimates *γ*_1_ = 7.885 (95% CI: 7.770, 8.008) and *γ*_2_ = 2.472 (95% CI: 2.299, 2.647) suggest that the extremely low CD4 counts are positively associated with AIDS progression and pre-AIDS death, while the former being more affected. Furthermore, for AIDS progression, the estimated shape parameter *α*_1_ = 2.627 (95% CI: 2.584, 2.686) is significantly greater than 1, suggesting the risk of AIDS progression increases gradually over time. In contrast, the risk of pre-AIDS death decreases over time with *α*_2_ = 0.797 (95% CI: 0.786, 0.810) significantly less than 1. The posterior mean of the spatial effect was estimated as 0.075, which indicates that 7.5% variation is accounted for by the spatial dependence.

Results capturing the longitudinal trajectories of all CD4 counts based on the log-Gaussian model are presented in Tables A.1 and A.2 (Appendix A.3), which demonstrate similar coefficient directions, further supporting the robustness of the primary findings.

### 4.2 Model evaluation

The spatiotemporal joint model demonstrates the best performance with the lowest DIC of *−*197,808.64, representing reductions of 2584.94, 102.70, and 2424.35 compared to the model with no random effects, the model removing either spatial effects, or temporal random effects, or both.

We evaluate the performance of survival model for AIDS progression using time-dependent ROC-AUC curves highlighting the advantage of our GP-based extreme downward excess longitudinal submodel. Compared with the common mean-based log-Gaussian model of CD4 counts (Figure 2), we see that the time-specified ROCs based on the GP-shared linear predictor are closer to the top-left corner. The time-dependent AUC values for the two competing risks joint models at different time cutoffs, along with the AUC curves considering extreme CD4 counts, consistently surpass those based on the mean, as illustrated in Figure 3. Furthermore, compared to the Gaussian model, the GP model demonstrates improved iAUC values, increasing from 0.7688 to 0.9183 for AIDS progression and from 0.6634 to 0.8901 for pre-AIDS death. Therefore, the GP-based extremely low CD4 counts demonstrate higher predictive power for HIV-AIDS progression and pre-AIDS death.

**Figure 2:**
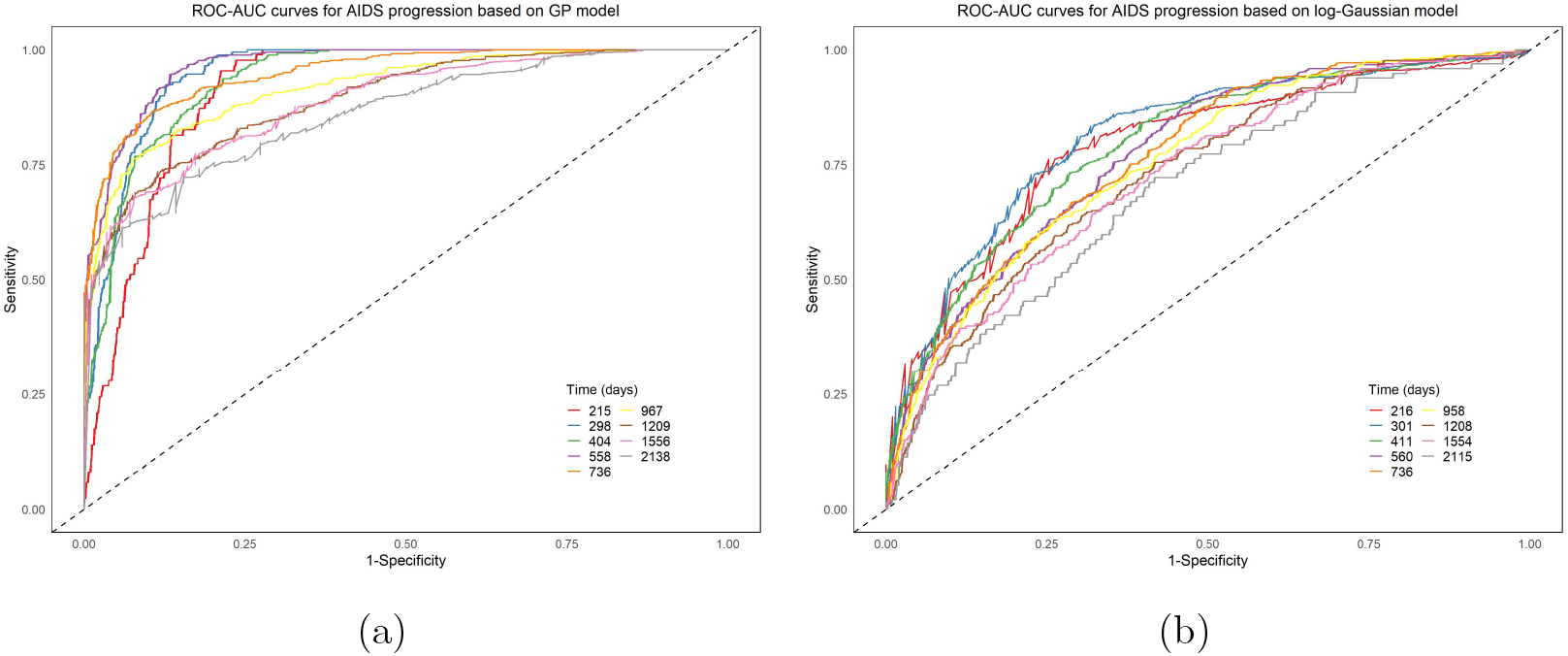
ROC-AUC curves for AIDS progression with shared GP submodel for extremely low CD4 counts on log scale (a) and log-Gaussian submodel for CD4 counts (b). Here the time slots are selected as deciles of the shared components from the 10% quantile being 215 and 216, and 90% quantile being 2138 and 2115 accordingly.

**Figure 3:**
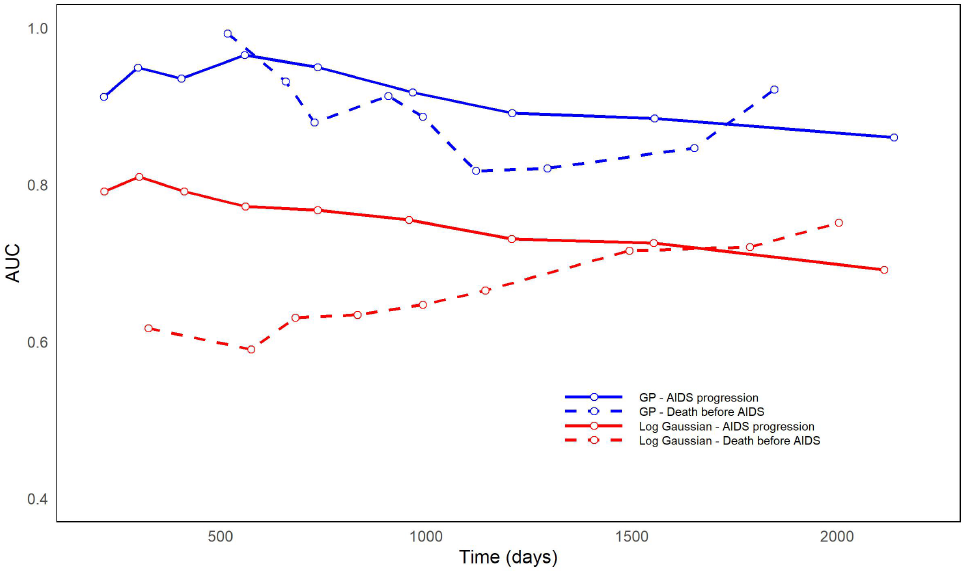
Time-dependent ROC-AUC curves for AIDS progression (solid line) and pre-AIDS death (dotted line) with shared GP submodel for extremely low CD4 counts on log scale in blue and log-Gaussian submodel for CD4 counts in red, with time slots selected using from 10% to 90% cutoffs.

## 5 Discussion

Joint longitudinal and survival modelling has become increasingly prevalent in biomedical research over the last decades (Ibrahim et al., 2010; Momenyan, 2021), allowing the exploitation of all available information from both types of data, thereby facilitating more efficient parameter estimation (Long and Mills, 2018). A key innovation of this study is the incorporation of extreme value theory into the downward excess of the CD4 longitudinal submodel and its shared effects in the two competing risks submodels, combined with the R-INLA implementation. Furthermore, the integrating spatial and temporal random effects in joint modelling provides new insights into the spatial cluster and temporal evolution of the disease progression over all counties in Zhejiang Province during the study period. This is a novel method of joint analysis that improves the accuracy of evaluating the predictor impact and offers a more precise and comprehensive approach to monitoring HIV-infected individuals and AIDS progression. The findings are expected to characterize the geographic inequalities in HIV/AIDS medical care and the survival status of patients with severe HIV, which are crucial for healthcare allocation, the development of effective surveillance programs and the improvement of targeted medical interventions.

Our study highlights the importance of monitoring the extreme decline in CD4 counts in HIV-infected individuals, providing a new perspective compared to previous studies that focused on mean CD4 counts (Dessiso and Goshu, 2017; Momenyan, 2021). We find that extremely low CD4 counts are more prevalent and prone to decline more sharply among heterosexual-infected patients, which is likely related to higher rates of late detection in this group. Larger European cohort studies have consistently identified heterosexual acquisition as a risk factor for delayed diagnosis (Ebrahim et al., 2013). Meanwhile, we see that the extremely low CD4 counts are generally lower in females compared to males and appear early in treatment, aligning with findings that females usually experience a more rapid decline in CD4 counts over a shorter period (Parsa et al., 2020). Furthermore, the significant impact of age at diagnosis and time between diagnosis and treatment indicates that older patients face more severe disease progression, especially with delayed treatment, corroborating previous research (Ford et al., 2015). Therefore, medical institutions should be prompted to strengthen the follow-up management of cases, increase the rate of antiviral treatment, and achieve early detection and treatment.

The survival quality for females is found to be lower than that for males, consistent with the findings from a related study (Ahmed et al., 2021). This disparity is likely to be influenced by traditionally restrictive attitudes for women in some rural areas of China, where the stigma and psychological stress associated with HIV/AIDS infection, combined with the physical discomfort of the disease, contribute to women experiencing lower survival quality compared to men (Sun et al., 2019). These social and cultural pressures further exacerbate the challenges women face in accessing timely healthcare and receiving adequate support, impacting their overall health outcomes. Homosexual transmission, on the other hand, acts as a protective factor in the disease progression of HIV/AIDS patients, with men who have sex with men (MSM) exhibiting greater awareness of HIV prevention and exhibiting more active testing behaviour (Patrick et al., 2017; Veinot et al., 2016). Furthermore, we observe that initiating ART 10 days earlier resulted in a reduction of 1.49% and 1.98% in the hazard ratios for AIDS progression and pre-AIDS death, respectively. Starting ART early helps prevent severe immune impairment, in line with the WHO’s 2016 recommendation for the rapid initiation of ART at the time of HIV diagnosis, regardless of CD4 count (Esber et al., 2020). For patients with extremely low CD4 counts, older HIV-infected individuals tend to have a lower risk of AIDS progression, which is consistent with research showing that late diagnosis of HIV (less than 12 months between diagnosis and AIDS onset) is more likely in individuals under 30 years of age (Schwarcz et al., 2006).

In our model, the estimated posterior mean of the spatial effect indicated that approximately 7.5% of the variation in AIDS progression and pre-AIDS death could be attributed to spatial dependence. This highlighted the importance of considering spatial factors when modelling HIV/AIDS dynamics and designing targeted interventions to address geographic differences in disease burden and healthcare accessibility (Laybohr Kamara et al., 2022; Zhang et al., 2017). For AIDS progression and pre-AIDS death, the shared regression coefficients of the longitudinal model were greater than 0, indicating that extremely low CD4 counts increased the risk of these events. The lower the initial CD4 count, the greater the impairment of the patient’s immune system function, leading to a poorer prognosis and increased risk of disease severity and mortality (Dorward et al., 2020; Shalev et al., 2020). This suggests that HIV/AIDS publicity should be intensified to accelerate the knowledge of HIV/AIDS, and the accessibility of testing services should be improved to encourage people with high-risk sexual behaviour to undergo HIV/AIDS testing as early as possible (Xu et al., 2021). Moreover, for AIDS progression, the estimated shape parameter in the Weibull model is significantly greater than 1, suggesting that the risk of AIDS progression increases gradually over time. In contrast, the risk of pre-AIDS death decreases over time with the shape parameter significantly less than 1. This decreasing risk is largely due to the fact that suicide is a major cause of pre-AIDS death progression (Park et al., 2022). A national cohort study in China found that suicide risk is 96 times higher than the general population in the first three months post-diagnosis (Chen et al., 2022), but this risk declines over time, contributing to the overall reduction in death risk before AIDS.

This joint modelling framework offers a general framework for capturing the progression of outcomes through extreme observations, making it applicable to diseases where extreme measurements are pivotal for diagnosis. For example, it could account for extreme variations in viral load in AIDS progression (Shoko and Chikobvu, 2019). Although our findings from Zhejiang Province in recent decades may not fully generalize to other periods or regions, due to the spatial disparities and varying healthcare systems across provinces (Xu et al., 2020), this framework still facilitates adjustment for spatial and temporal clustering, enhancing its adaptability for large-scale studies across different regions. Additionally, the unknown adherence to treatment among patients may affect the capture of changes in CD4 counts over time in the longitudinal data within the model and hinder accurate interpretation of the relationship between CD4 dynamics and clinical outcomes (Barnes et al., 2020). Future studies should consider estimating such uncertainties.

## 6 Conclusion

This paper assesses the impact of multiple clinical predictors and extremely low CD4 counts on AIDS progression and pre-AIDS death using R-INLA. The joint extreme longitudinal and competing risks models enable us to reveal the pivotal factors, including the demographic factors, route of infection, and diagnosis-relevant factors in HIV/AIDS risk. The findings have significant implications for the development of effective evidence-based strategies to mitigate the impact of HIV/AIDS. Through joint analyses, we can gain a more comprehensive understanding of the process of AIDS progression, elucidate the underlying drivers of the trends and associations we observed, and provide critical information for policy development and resource allocation. This will support the design of targeted and effective interventions to better control the spread of HIV/AIDS in the province and promote public health and social well-being.

## Data Availability

Data are available from the authors upon reasonable request and with permission of the Zhejiang Provincial Center for Disease Control and Prevention.

## A Appendix

## A.1 Peaks-over-threshold model

Peaks-over-threshold (POT) characterizes the threshold exceedance with the generalized Pareto (GP) model (Opitz et al., 2018; Spearing et al., 2023). Given a random variable *X* with distribution *F* (*x*) and a large threshold *u* approaching *x*^*\**^ = sup*{x* : *F* (*x*) *<* 1*}* (the right end-point), the distribution of threshold excess can be approximated by the GP distribution,

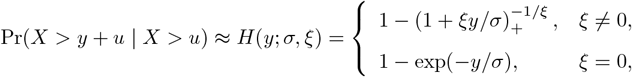

where *x*_+_ = max(*x*, 0) and scale parameter *σ* = *σ*(*u*) *>* 0 depends on the threshold *u*. The shape parameter *ξ ∈* ℝ determines the tail behavior. Specifically, the case with *ξ >* 0, *ξ <* 0, and *ξ* = 0 corresponds to the risk with a heavy tail, light tail, and exponential tail, accordingly.

Note that for our interest in the extremely low log (CD4), selecting the lower threshold *u* for *y*_*ij*_(*s, t*) is equivalent to selecting the upper threshold *u*^*\**^ = *−u* for the *−y*_*ij*_(*s, t*). We followed the rule of thumb (Haan and Ferreira, 2006) and selected the 15% quantile *u* = 5.7 of log (CD4) as the threshold. Namely, CD4 counts below exp(5.7) = 299 cells*/*mm^3^ are considered extremely low in our context.

Moreover, we use the reparametrization of GP distribution (Opitz et al., 2018) with the tail index *ξ* and a specific *q*-th quantile *κ*_*q*_ for some fixed probability of interest *q ∈* (0, 1),

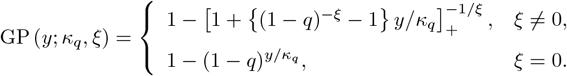

This reparametrization avoids problems due to the correlation among estimated GP parameters in INLA. In particular, we use *q* = 0.6 in our research.

### A.2 Joint model based on log-Gaussian distribution

Let *y*(*t*) denote the logarithmic transformation of the CD4 count at day *t* since the HIV-infected, following a Gaussian law in the longitudinal submodel. To be specific, we suppose that

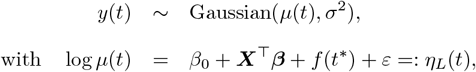

where the location parameter *μ*(*t*) varies over time and the scale parameter *σ* remains constant (Pan et al., 2024). In the survival submodel, we consider the same cause-specific hazard functions as those presented in Section 3.2. The scale parameter is modeled using a log-linear component with mixed effects, as specified in Eq.(3.3). Both the longitudinal and survival submodels specify the random effects, fixed effects, and error terms, the same as for the model described in detail in Sections 3.1 and 3.2.

### A.3 Model results based on log-Gaussian longitudinal submodel

**Table A.1:** Summary of posterior estimates of fixed effect in the joint model fitted by competing risks (AIDS progression and pre-AIDS death) and longitudinal measurements (CD4 counts), based on log-Gaussian distribution.

| **Longitudinal endpoint: CD4 count** |  |  |  |
| --- | --- | --- | --- |
| **Predictors** | Mean | 2.5% quantile | 97.5% quantile |
| **Gender** |  |  |  |
| Female | Reference |  |  |
| Male | 0.006 | −0.012 | 0.025 |
| **Marital status** |  |  |  |
| Single | Reference |  |  |
| Married | −0.006 | −0.021 | 0.007 |
| **Route of infection** |  |  |  |
| Heterosexual transmission | Reference |  |  |
| Homosexual transmission | −0.013 | −0.028 | 0.002 |
| **Age at diagnosis (in $10^{-1}$)** | −0.009 | −0.014 | −0.004 |
| **Treatment delay (in $10^{-3}$)** | 0.073 | −0.042 | 0.188 |
| **Survival endpoint 1: AIDS progression** |  |  |  |
| **Predictors** | Mean | 2.5% quantile | 97.5% quantile |
| **Gender** |  |  |  |
| Female | Reference |  |  |
| Male | −2.399 | −2.553 | −2.245 |
| **Marital status** |  |  |  |
| Single | Reference |  |  |
| Married | −0.207 | −0.357 | −0.057 |
| **Route of infection** |  |  |  |
| Heterosexual transmission | Reference |  |  |
| Homosexual transmission | −1.535 | −1.708 | −1.362 |
| **Age at diagnosis** | −0.051 | −0.056 | −0.045 |
| **Treatment delay** | −0.014 | −0.016 | −0.013 |
| **Survival endpoint 2: Pre-AIDS death** |  |  |  |
| **Predictors** | Mean | 2.5% quantile | 97.5% quantile |
| **Gender** |  |  |  |
| Female | Reference |  |  |
| Male | −8.526 | −9.025 | −8.027 |
| **Marital status** |  |  |  |
| Single | Reference |  |  |
| Married | −7.353 | −1.136 | −0.335 |
| **Route of infection** |  |  |  |
| Heterosexual transmission | Reference |  |  |
| Homosexual transmission | −7.111 | −7.716 | −6.506 |
| **Age at diagnosis** | −0.026 | −0.040 | −0.011 |
| **Treatment delay** | −0.014 | −0.016 | −0.013 |

**Table A.2:** Summary of the posterior estimates of the parameters, based on log-Gaussian distribution.

| Parameter | Mean | 2.5% quantile | 97.5% quantile |
| --- | --- | --- | --- |
| Shape parameter of AIDS progression ($\alpha_1$) | 0.802 | 0.801 | 0.803 |
| Shape parameter of pre-AIDS death ($\alpha_2$) | 0.797 | 0.786 | 0.810 |
| BYM2 mixing parameter ($\phi$) | 0.042 | 0.041 | 0.043 |
| Shared scale of AIDS progression ($\gamma_1$) | -1.138 | -1.141 | -1.136 |
| Shared scale of pre-AIDS death ($\gamma_2$) | -0.556 | -0.558 | -0.553 |

## Declaration

## Ethics approval and consent to participate

This study is conducted in accordance with the ethical principles of the Declaration of Helsinki and is approved by the Institutional Review Board of the School of Public Health, Fudan University (#2014-03-0497).

## Consent for publication

Not applicable.

## Competing interests

The authors declare that they have no competing interests.

## Funding

This research was partially supported by the National Natural Science Foundation of China (71473046), the Research Development Fund (RDF1912017), and the Post-graduate Research Fund (PGRS2112022) at Xi’an Jiaotong-Liverpool University.

## Authors’ contributions

C.L. and Y.T. developed the methodology and conceptualization. Y.T. prepared the initial draft of the paper. Y.T. and K.W. programmed the code. C.L., H.Z., and K.W. analyzed the main results. Y.C. and W.C. processed the original data. N.H., Y.W., R.K.D., L.C., C.L., and W.C. commented on and revised manuscript drafts. All authors read and approved the final manuscript.

## Acknowledgements

We thank the editors and anonymous reviewers for their helpful remarks.

## Notes

### Competing Interest Statement

The authors have declared no competing interest.

### Author Declarations

This study is conducted in accordance with the ethical principles of the Declaration of Helsinki and is approved by the Institutional Review Board of the School of Public Health, Fudan University (\#2014-03-0497).

